# Analysis of Adverse Drug Events of Opioids in the United States

**DOI:** 10.1101/2022.10.21.22281281

**Authors:** Edward Y. Liu, Kenneth L. McCall, Brian J. Piper

## Abstract

The United States (US) is going through an opioid crisis with annual increases in opioid-related mortality. Our study analyzed the adverse drugs events (ADEs) for eleven prescription opioids when correcting for distribution, and their ratios for three periods: 2006-2010, 2011-2016, and 2017-2021 in the US. The opioids included buprenorphine, codeine, fentanyl, hydrocodone, hydromorphone, meperidine, methadone, morphine, oxycodone, oxymorphone, and tapentadol. The Food and Drug Administration Adverse Event Reporting System (FAERS) database consists of reports by MedWatch adverse event forms submitted by healthcare professionals and others (N=667,969), whereas the Automation of Reports and Consolidated Orders System (ARCOS) reports on medically used controlled substances. Oral morphine milligram equivalents (MMEs) were calculated by conversion relative to morphine. The relative ADEs of the select opioids calculated from FAERs, opioid distribution from ARCOS, and the FAERs to ARCOS ratios were analyzed for the eleven opioids. Oxycodone reports peaked in the third period and showed consistently high ADEs. Codeine and meperidine accounted less than five percent of ADEs. The ARCOS distributions were relatively constant over time, but methadone consistently accounted for the largest portion of the total distribution. The FAERS to ARCOS ratios generally increased over time, with meperidine (60.6), oxymorphone (11.1), tapentadol 10.3, and hydromorphone (7.9) most over-represented for ADEs in the third period. Oxymorphone had a 542.2% increase in ratio between the second and third period should be noted. Methadone was under-represented (< .20) in all three periods. These findings indicate the need to further monitor and address the ADEs of select opioids.

**Significance:** Prescription opioid use in the US is among the highest in the world. This study analyzed both FAERs and ARCOS databases to understand the adverse drugs events. This investigation identified which opioids were overrepresented (e.g. meperidine) and underrepresented (e.g. methadone) for adverse effects relative to the prevalence of use to inform healthcare policies and change the way physicians view and prescribe these opioids.

## Introduction

The opioid epidemic in the United States (US) has escalated to a nationwide public health crisis due to annual elevations in opioid-related mortality (Lyden et al., 2019; O’Donnell et al., 2017; Singh et al., 2019). Drug overdose deaths increased by 4-fold from 1999 to 2017 with opioid-related deaths accounting for two-thirds of the deaths (Singh et al., 2019). Death rates related to opioid overdose increased by 27.9% from 2015 to 2016 (Seth, 2018). The CDC has recently indicated that the number of drug overdose deaths increased by nearly 5% from 2018 to 2019, with over 70% of the 70,630 drug-related deaths in 2019 involving opioids (Centers for Disease Control and Prevention, 2021). Although the volume of opioids prescribed in the US decreased from 2010 to 2015 after peak in 2011, the amount is still significantly higher relative to 1999 (Guy, 2017; Mack et al., 2018; Piper et al., 2018). An analysis of the International Narcotics Control Board records from 2015 to 2017 revealed that 10% of the world’s population consumed 89% of the world’s supply of prescription opioids. Further, the US ranked third for highest opioid consumption per capita (Richards et al., 2022).

A recent study examining the national trends in opioid exposures reported to US poison control centers indicated that the proportion of exposures with adverse drug events (ADEs) increased despite the overall decrease in frequency and rate of opioid exposures from 2011-2018 (Rege et al., 2021). ADEs are reported in the US Food and Drug Administration Adverse Event Reporting System (FAERS), a large government database that consists of ADEs and medication error reports submitted through the MedWatch Program primarily from healthcare professionals (Zhou & Hultgren 2020). In addition to using the FAERS database to quantify the adverse effects, we used the Drug Enforcement Administration’s Automation of Reports and Consolidated Orders System (ARCOS), a comprehensive data collection system where Schedule II and III controlled substances are mandatorily reported when distributed to pharmacies, hospitals, Narcotic Treatment Programs (NTP), and long-term care facilities (Piper et al., 2018; U.S. Drug Enforcement Administration, 2022). We used both databases to identify the ADEs of several common Schedule II and III prescription opioids relative to their distributions in the US for the past one and a half decades. This analysis identifies which opioids were over, or under, represented for ADEs relative to their use.

## Methods

### Procedures

The FDA FAERS and ARCOS databases were queried from 2006 to 2021 to examine the ADEs and distribution of opioids, respectively of the eleven opioids. These opioids were selected based on previous studies and include nine for pain: codeine, fentanyl, hydrocodone, hydromorphone, meperidine, methadone, morphine, oxycodone, oxymorphone, and tapentadol, and two mainly used for opioid use disorders (OUD): buprenorphine and methadone (Cabrera et al., 2019; Eidbo et al., 2022; Mack et al. 2018; Modarai et al., 2013; Singh et al, 2019; Veronin et al., 2019). The search involved only generic opioid names indicated in the FAERS database with ADEs including misuses, overdoses, serious cases, and deaths (U.S. Food and Drug Administration 2022). Procedures were approved as exempt by the IRB of Geisinger and the University of New England.

### Statistical Analysis

The total oral morphine milligram equivalent (MME) was calculated based on the weight for all eleven opioids and expressed in three periods (2006 to 2010; 2011 to 2016; 2017 to 2021) for the US, excluding the US territories. Hereafter, these periods are referred to the first, second, and third, respectively. The first period was of increases in prescription opioid distribution, 2011 was the peak year, and the third period was of further declines in opioids used for pain and an escalation in OUD treatment (Collins et al., 2019; Pashmineh et al. 2020; Piper et al., 2018). The top three reaction group and reactions were reported for each opioid from 2006-2021 with percentages indicating the amount relative to the number of adverse events within that period. Three analyses were also completed for each period: (1) the frequency of ADEs of each opioid based on FAERS, (2) the percentage of total opioid distribution based on ARCOS, and (3) the FAERS to ARCOS ratios. The oral MME was calculated to correct for the relative potency of each opioid relative to morphine. The conversions were as follows: buprenorphine (10), codeine (0.15), fentanyl base (75), hydrocodone (1), hydromorphone (4), meperidine (0.1), methadone (10), morphine (1), oxycodone (1.5), oxymorphone (3), and tapentadol (0.4) (Eidbo et al., 2022; Piper et al., 2018). We identified any ratio >1.0 as an overrepresentation and <1.0 as underrepresentation of the ADEs of the opioid when correcting for distribution. For example, an opioid which accounted for 10% of ADEs but 5% of distribution would have a ratio of 2.0 (i.e. over-represented). Data analysis and figure preparation were completed with GraphPad Prism, version 9.3.1.

## Results

We queried data from FAERS and ARCOS databases for the eleven opioids from 2006 to 2021. Supplementary Figure 1 indicates the percentage of ADEs reports submitted by healthcare professionals to FAERs for the eleven opioids from 2006-2021. Almost one-third (31.2%) of the reports were from these individuals. Codeine (72.9%), meperidine (70.5%), and methadone (68.1%) had the most submissions from healthcare workers, while the remaining eight opioids ADEs were acquired mainly from patients. Table 1 indicates the ADEs reports from 2006-2021 obtained from FAERs. Oxycodone, fentanyl, and morphine were responsible for over half (55.2%) of the total number of ADEs (N=667,969) while meperidine accounted for less than one percent. The top three most common reaction groups included: injury, poisoning, and procedural complications; general disorders and administration site conditions; and psychiatric disorders. Common specific reactions consisted of abnormal drug effects (e.g. dependence, hypersensitivity, ineffectiveness), overdose, and death. The death rates varied among the opioids with oxymorphone having the largest proportion from death as “reaction” (40.6%) and death as “outcome” (70.6%), while meperidine had the least with 1.4% and 7.4%, respectively (Table 2). Figure 1 shows the percentage of ADEs for each opioid. The opioids fell into three groups in 2017-2021, high (>15%): oxycodone and morphine; intermediate (5-15%): hydromorphone, fentanyl, buprenorphine, hydrocodone, oxymorphone, tapentadol; and low (<5%): methadone, codeine, and meperidine. Oxycodone was consistently high across all periods: 2006-2010 (19.9%), 2011-2016 (17.8%), and 2017-2021 (26.0%). Fentanyl accounted for the largest portion of ADEs in the first two periods (2006-2010 (41.6%), 2011-2016 (23.6%)), but decreased greatly since the second (−49.9%). Methadone showed a noticeable decrease (−55.8%) from the second to third period, while oxymorphone indicated a marginal (+114.5%) increase. Codeine and meperidine accounted for less than five percent of the total ADE reports in all periods.

**Table 1.**
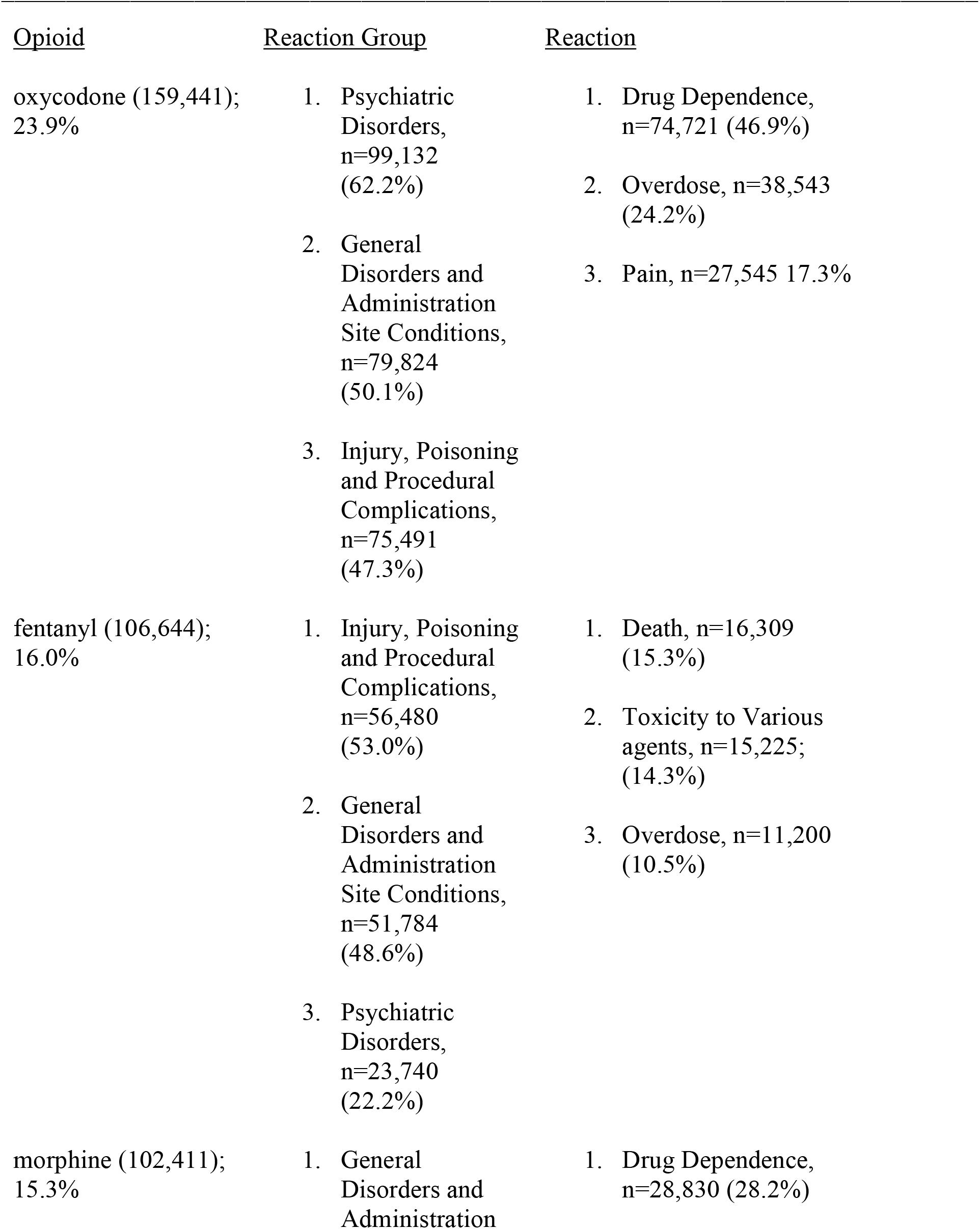
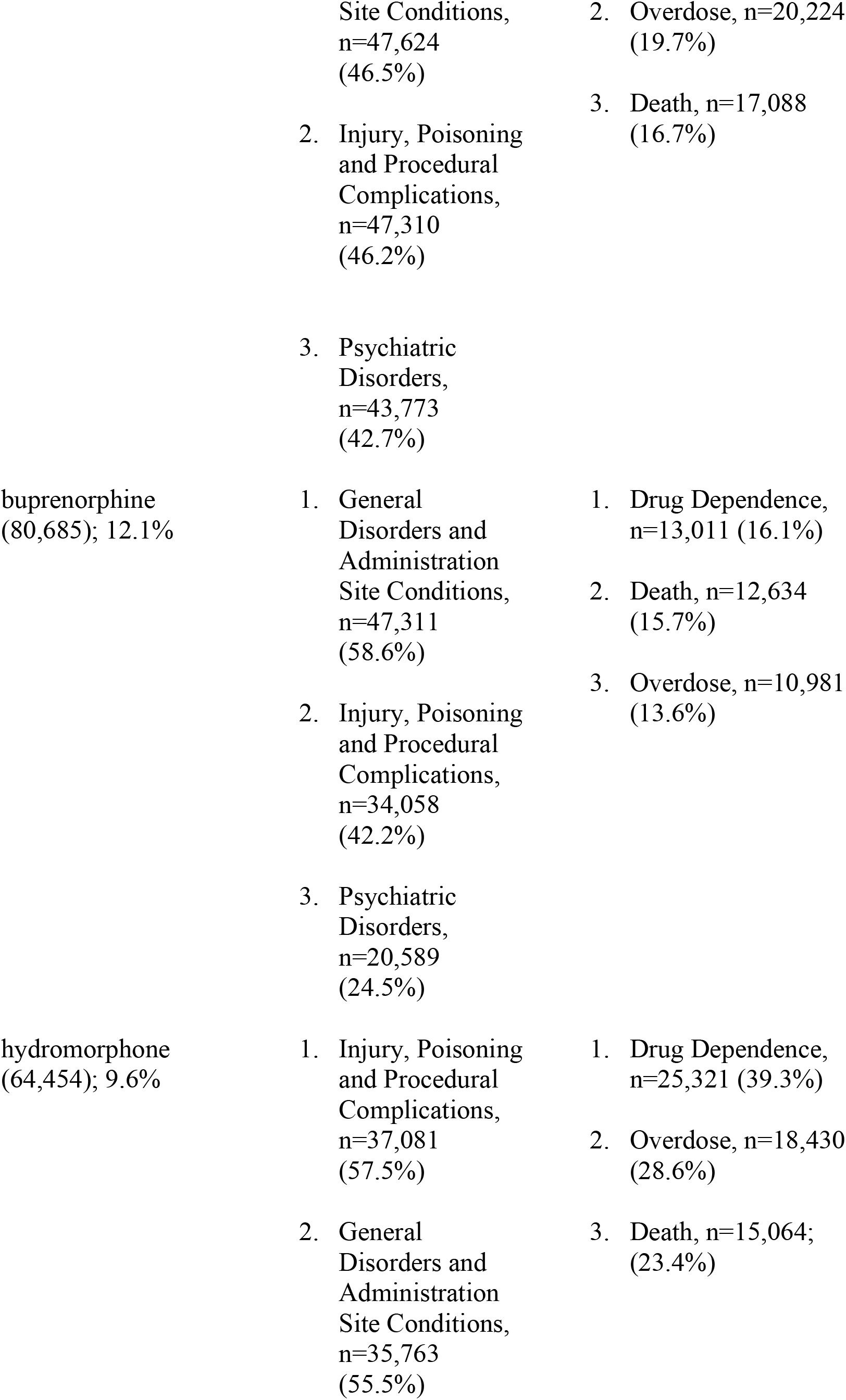
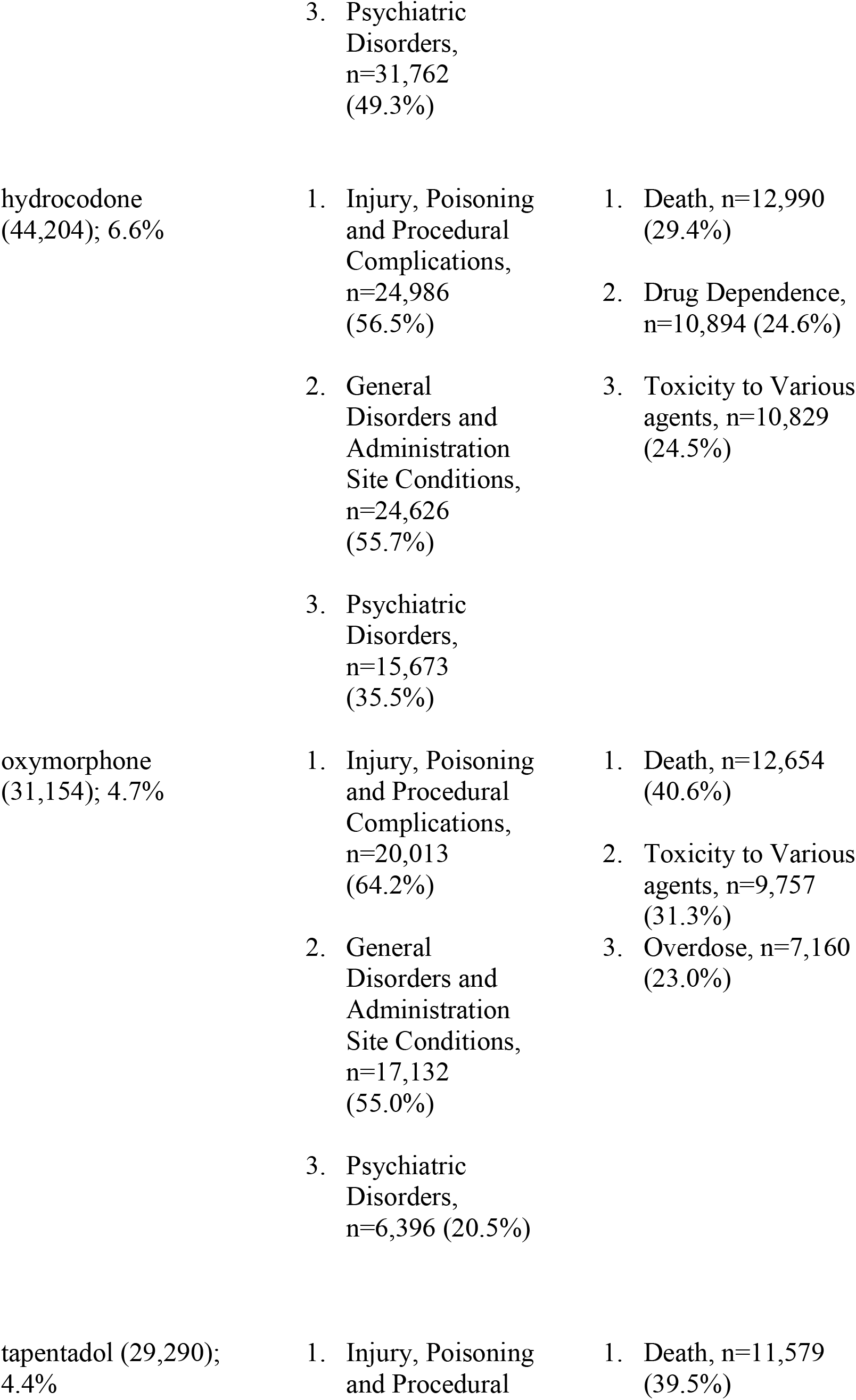
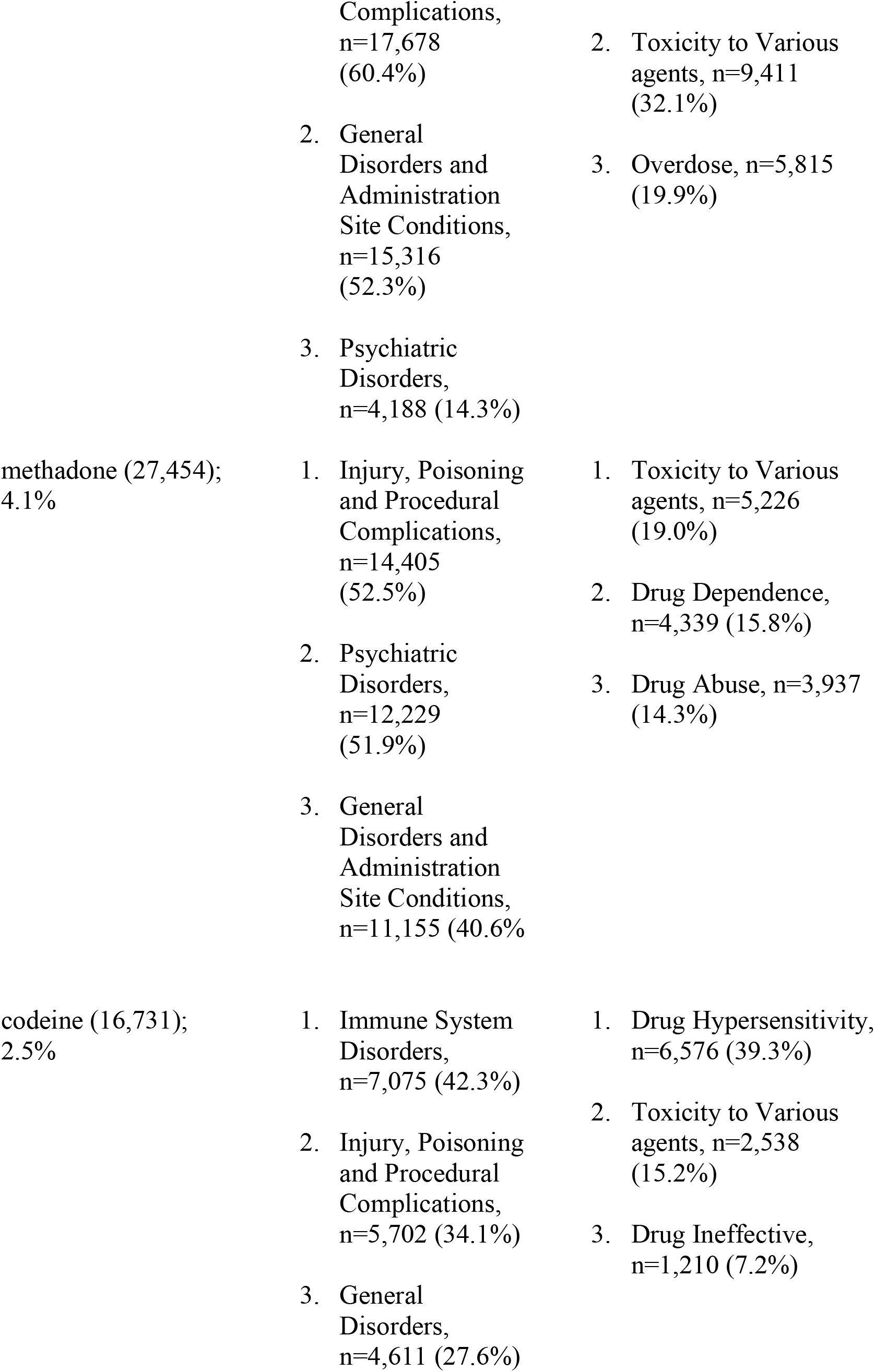
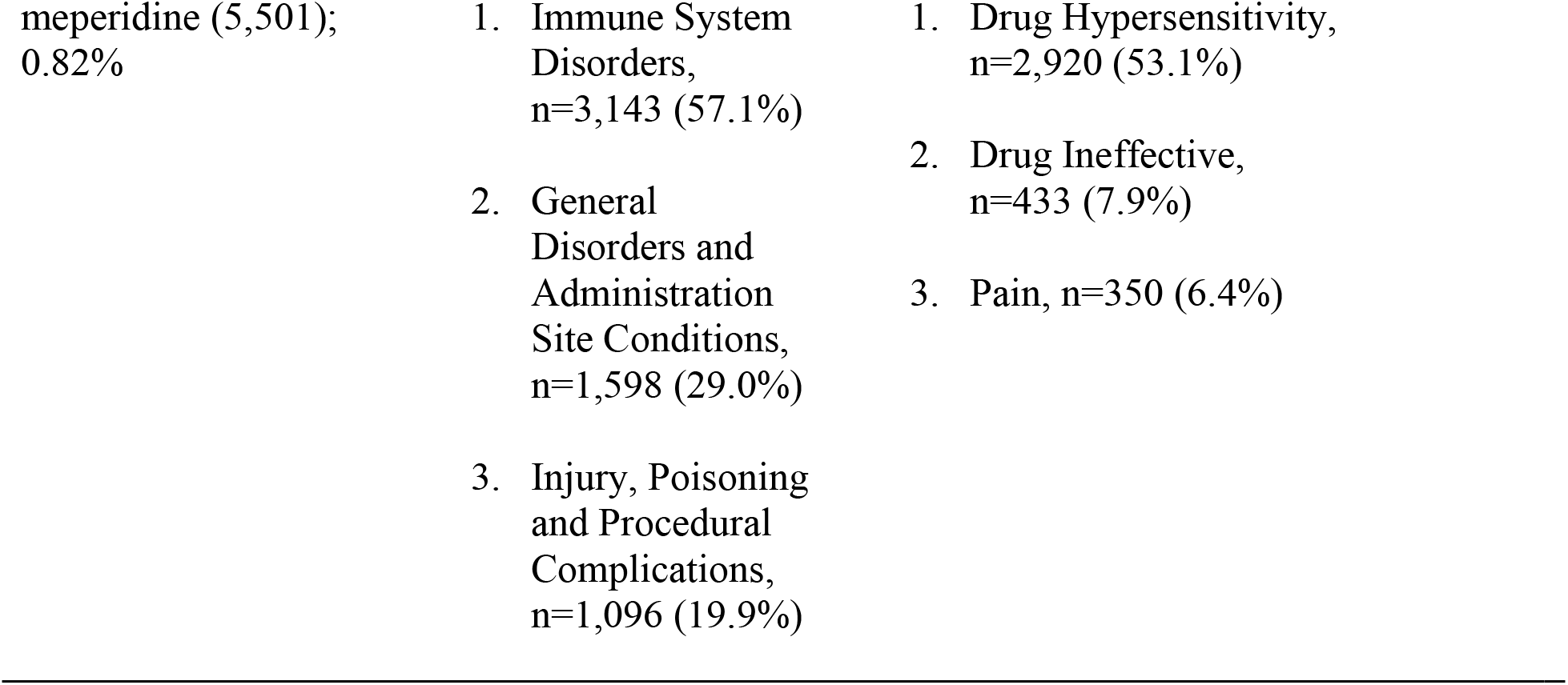
Adverse event reports by count and percentage in the US Food and Drug Administration’s Adverse Effect Reporting System for eleven prescription opioids for 2006-2021. The three most common reaction groups and reactions are shown.

**Table 2.**
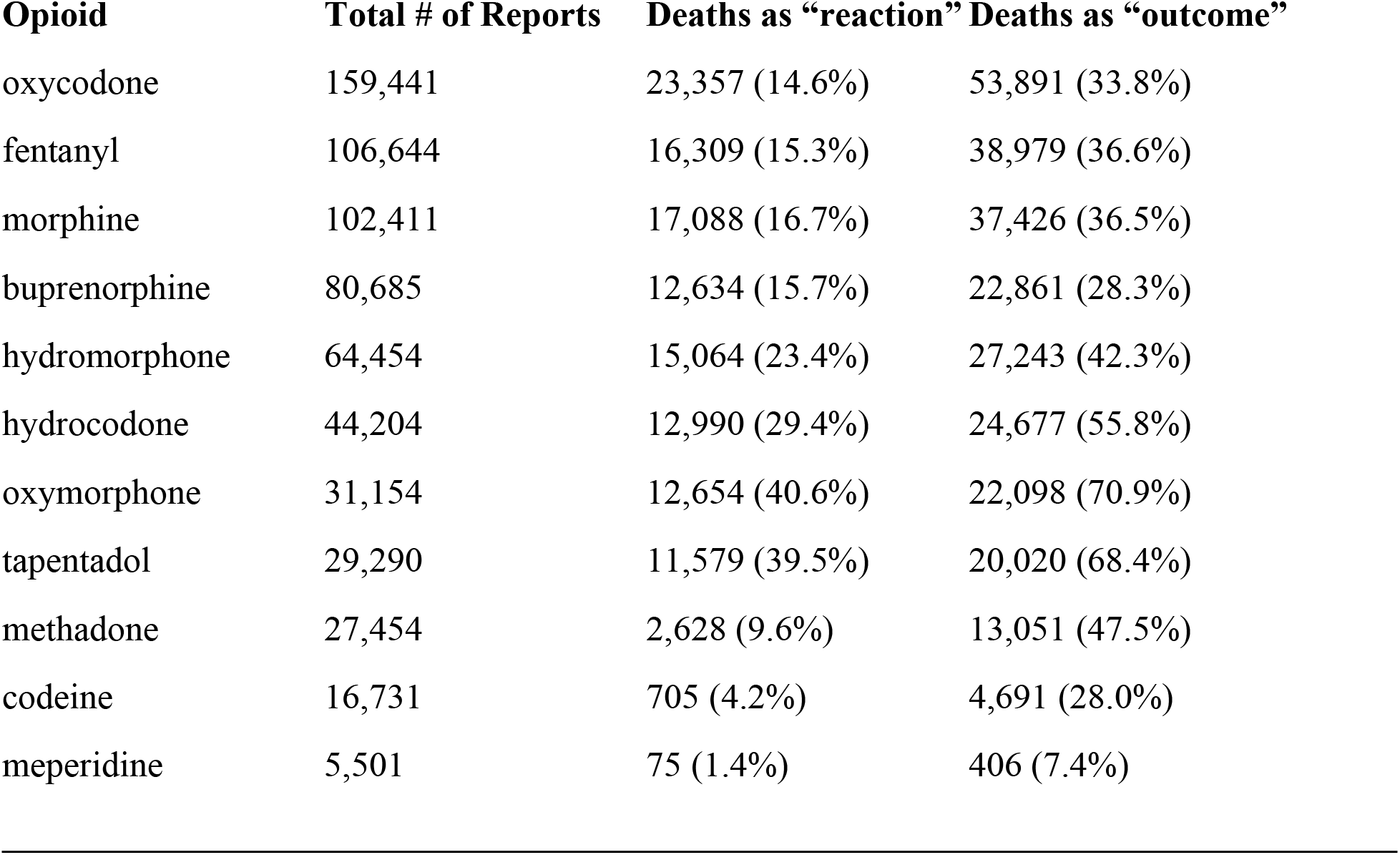
Death reports and their relative percentages by “reaction” and “outcome” in the US Food and Drug Administration’s Adverse Effect Reporting System for eleven prescription opioids for 2006-2021.

| Opioid | Total # of Reports | Deaths as “reaction” | Deaths as “outcome” |
| --- | --- | --- | --- |
| oxycodone | 159,441 | 23,357 (14.6%) | 53,891 (33.8%) |
| fentanyl | 106,644 | 16,309 (15.3%) | 38,979 (36.6%) |
| morphine | 102,411 | 17,088 (16.7%) | 37,426 (36.5%) |
| buprenorphine | 80,685 | 12,634 (15.7%) | 22,861 (28.3%) |
| hydromorphone | 64,454 | 15,064 (23.4%) | 27,243 (42.3%) |
| hydrocodone | 44,204 | 12,990 (29.4%) | 24,677 (55.8%) |
| oxymorphone | 31,154 | 12,654 (40.6%) | 22,098 (70.9%) |
| tapentadol | 29,290 | 11,579 (39.5%) | 20,020 (68.4%) |
| methadone | 27,454 | 2,628 (9.6%) | 13,051 (47.5%) |
| codeine | 16,731 | 705 (4.2%) | 4,691 (28.0%) |
| meperidine | 5,501 | 75 (1.4%) | 406 (7.4%) |

**Figure 1.**
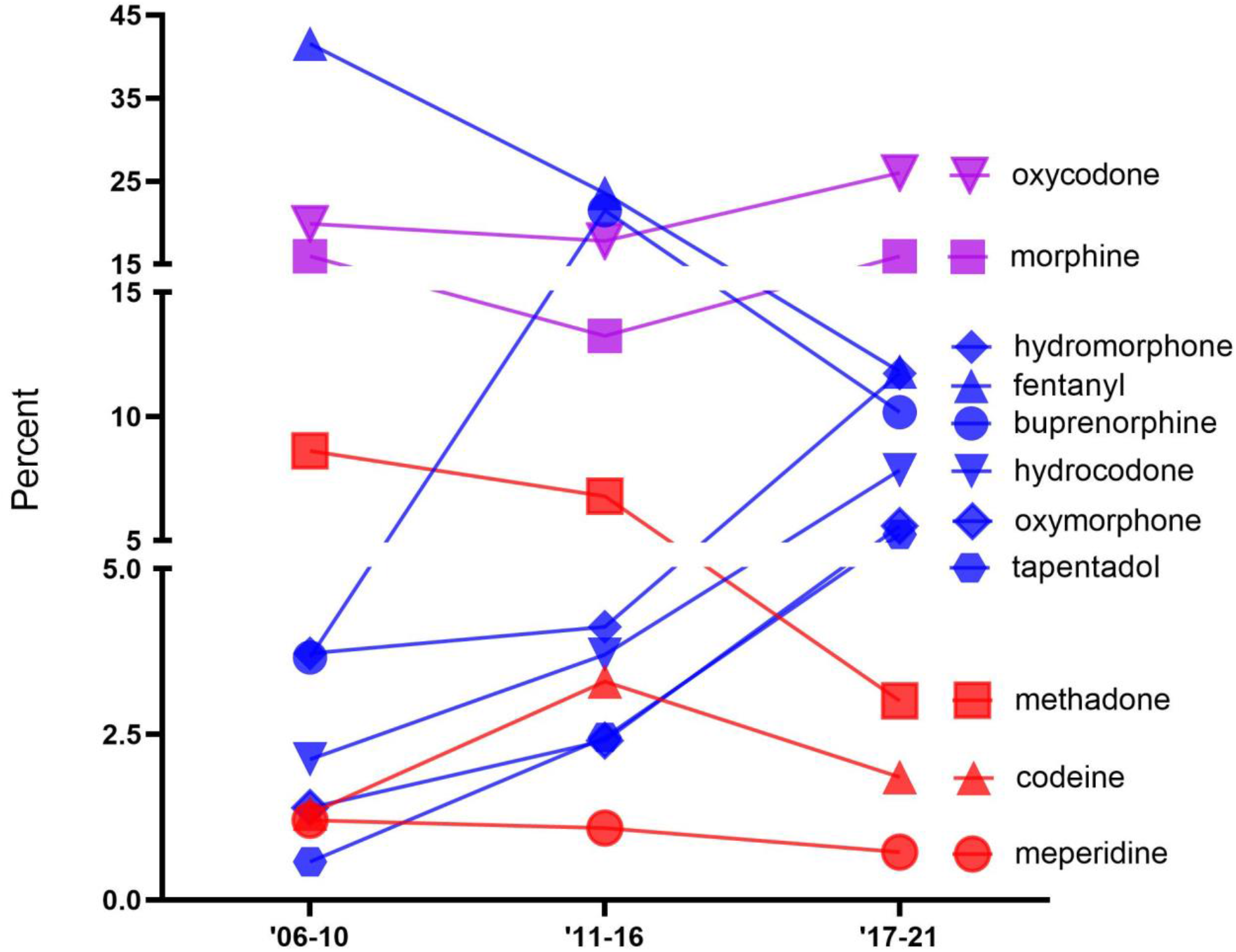
Percent of adverse drugs events (ADEs) of the eleven prescription opioids obtained from the FDA Adverse Event Reporting System over time.

Figure 2 depicts the percent of the total MME due to each opioid over time. The opioids fell into three groups which were generally stable over time: high: methadone and oxycodone; intermediate: buprenorphine, hydrocodone, morphine, and fentanyl, and low: hydromorphone, codeine, oxymorphone, tapentadol, and meperidine. Methadone accounted for two-fifths to one-half of the total distribution: 2006-2010 (44.2%), 2011-2016 (40.2%), and 2017-2021 (47.3%). Codeine, tapentadol, and meperidine consistently made up less than one percent of distribution.

**Figure 2.**
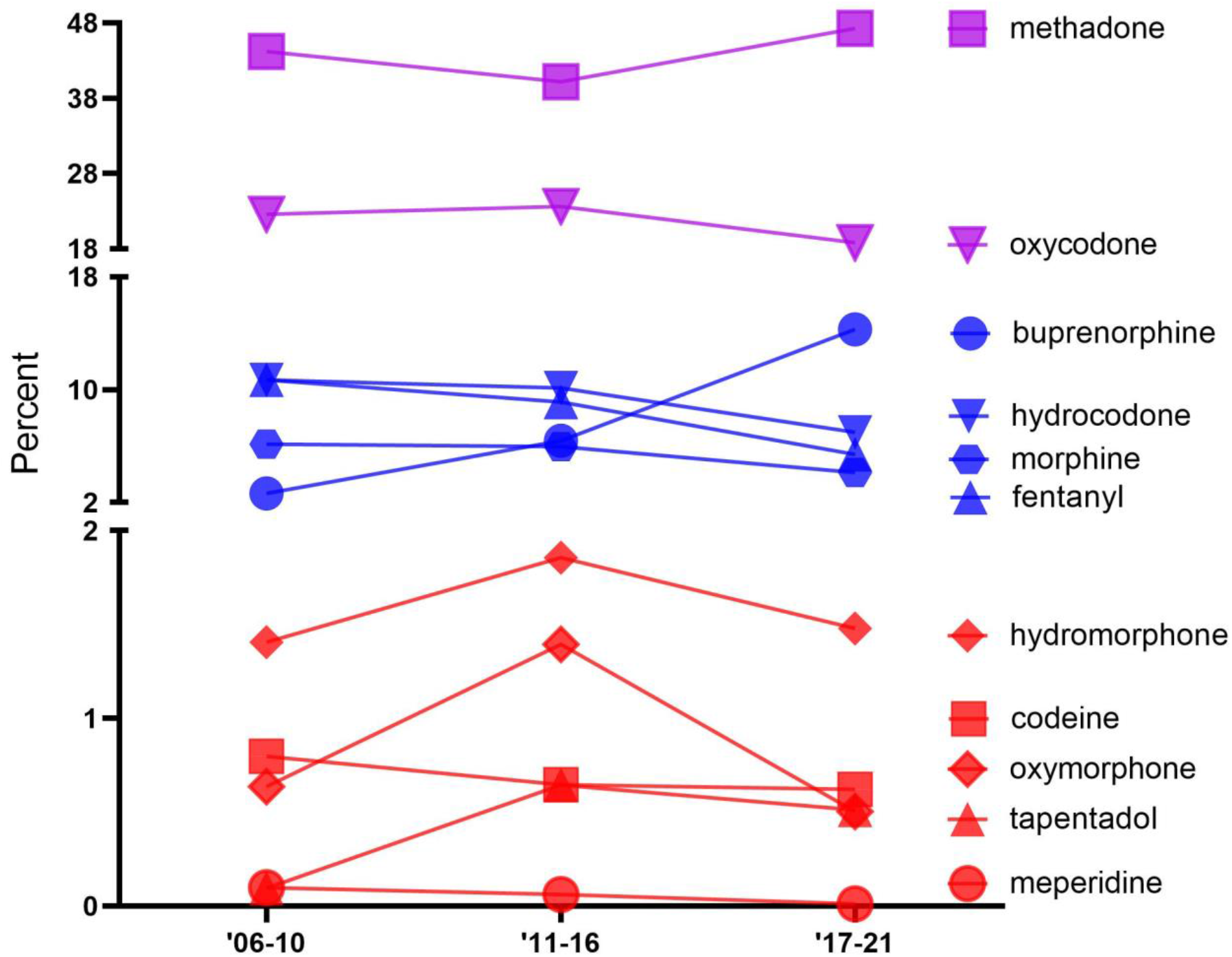
Percent of the total morphine mg equivalent (MME) of the distribution of eleven prescription opioids as reported to the Drug Enforcement Administration’s Automated Reports and Consolidated Orders System (ARCOS) over time.

Figure 3 shows the ADE to distribution ratio for each opioid for each period. Most drugs indicated an overrepresentation in the FAERs to ARCOS proportion. The general pattern showed increases over time with the most over-represented in the third period being meperidine (60.6) followed by oxymorphone (11.1), tapentadol (10.3), and hydromorphone (7.9). Oxymorphone showed the largest increase (+542.2%) in proportion from the second to third period followed by hydromorphone (+257.7%) and meperidine (+245.2%). Buprenorphine indicated the greatest decrease (−371.3%) followed by codeine (−71.0%), and fentanyl (−18.2%). Methadone was under-represented (< .20) in the three periods.

**Figure 3.**
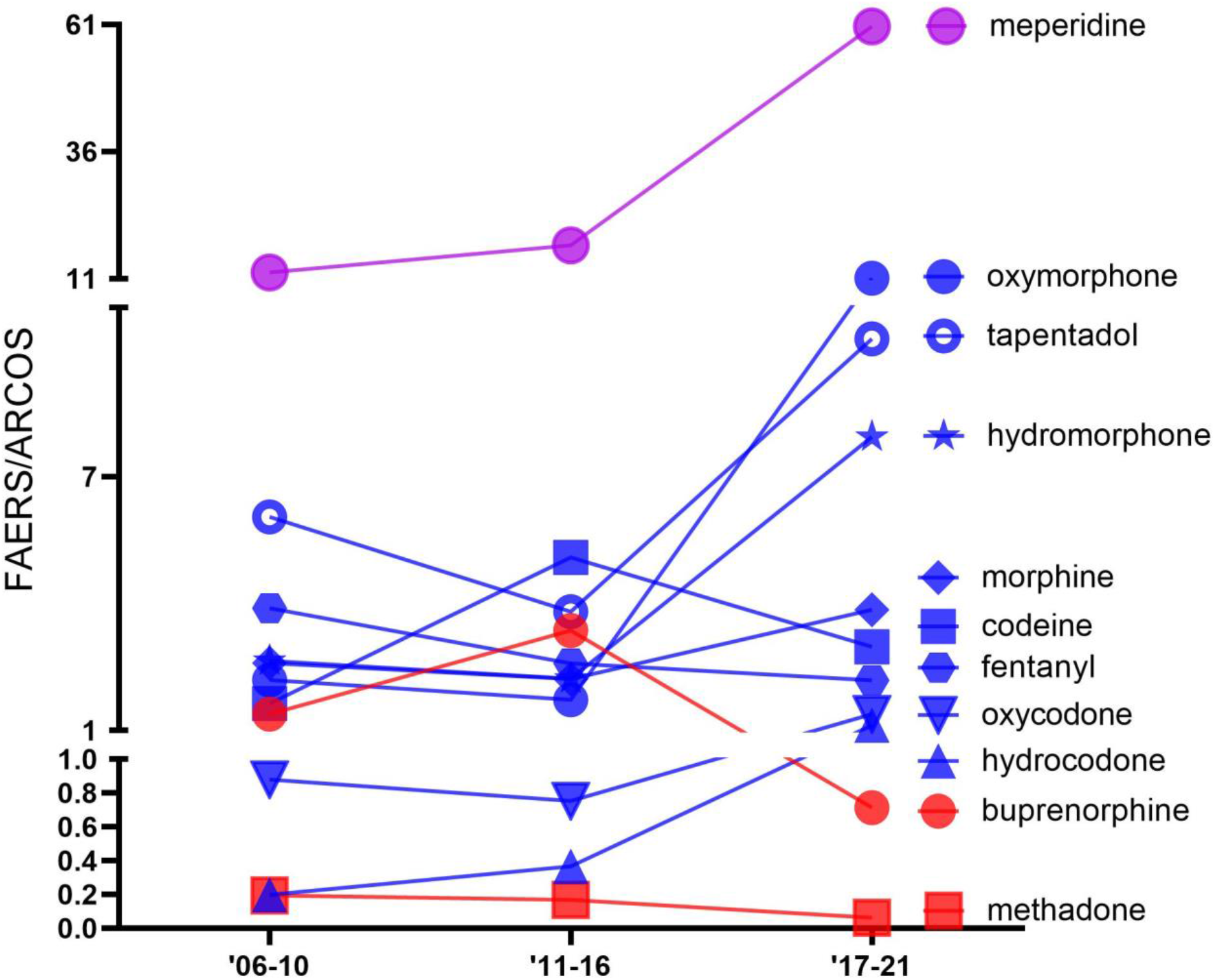
Ratio of the percent of adverse drug events as reported to the US Food and Drug Administration’s Adverse Events Reporting System relative to the percent of distribution as reported to the Drug Enforcement Administration’s Automated Reports and Consolidated Orders System databases for eleven prescription opioids over time. Values greater than 1.0 are over-represented and values less than 1.0 are under-represented.

## Discussion

Our study demonstrated varied ADEs for eleven commonly used opioids. This is also the first report to describe the ADEs while correcting for the prevalence of each opioids US distribution.

Oxycodone, fentanyl, and morphine made up over half (55.2%) of the total number of ADEs (N=667,969) with meperidine composing less than one percent. Oxycodone has a high potential for misuse due to its high reinforcing characteristics and its administration methods, including pill crushing for immediate release and through IV injections, leading to high dependence (Kibaly et al., 2020; Table 1). Fentanyl with its high potency and abuse potential, tendency to be mixed with other drugs, and illegal distribution may contribute to the high ADEs across all three periods (Comer & Cahill, 2019; DEA, 2016; National Institute on Drug Abuse, 2021). Like other opioids, morphine tolerance can develop secondary to continuous usage due to changes in receptor density and G protein coupling receptors and its signal transduction pathway (Listos et al., 2019). Meperidine with its consistently low (<1%) relative distributions of ADEs may be secondary to its ability to cause lethal ADEs including serotonin syndrome and psychological or physical dependence (Boyle et al., 2021). The distribution of this once ubiquitous agent has continued to decline (Harrison et al., 2022).

Methadone and buprenorphine both showed increases in counts in the past decade mostly due to their use for OUD with buprenorphine specifically showing a 122.5% increase in hospital consumption in the past decade (Bishop-Freeman et al., 2021; Cicero et al., 2014; Eidbo et al., 2022; Furst et al., 2022; Mattick et al., 2014; Pashmineh et al., 2020). These opioids have been commonly used to treat opioid dependence, and with more Medicaid coverage, their prevalence has been rising (Burns et al., 2016; Mattick et al., 2014). The high counts of oxycodone may be attributed to its common use and effectiveness for treating moderate-to severe acute pain (Davis et al., 2021; Moradi et al., 2012).

Given that meperidine demonstrated the lowest frequency of ADEs, it was surprising to find that its adverse effects were the most overly represented compared to its distribution (60.6), particularly in the third period (60.6). In contrast to Veronin et al. (2019) indicating oxycodone had high death to count percentages compared to other opioids, ours indicated oxycodone’s hovered around 1% of the relative ratios throughout as seen in figure 3. The decline in oxycodone overdoses might be attributed to the reduction in abuse since the development of its extended release in late 2010 (Johnson et al., 2014). Oxymorphone’s notable increase in adverse effects (+542.2%) relative to its counts was unsurprising, as it constantly had low counts throughout the three periods relative to other opioids. Oxymorphone as a schedule II drug has high potency and misuse potential related to its euphoric effects explaining the huge increase in proportion which might also explain its high proportion of deaths from ADEs (DEA 2019). It also tapentadol with its dual mechanism of action acting as both a μ-opioid receptor agonist and noradrenaline reuptake inhibitor has indicated better tolerability than other commonly prescribed opioids due to its low μ-load (Romualdi et al., 2019). It was therefore unexpected to see its overrepresentation (10.3) in the third period. Furthermore, hydromorphone’s pronounced decrease in distribution in the past decade in addition to its high potential for fatality and overdose rates may contribute to its overrepresentation since the second period (+257.7%) (Eidbo, et al., 2022; Lowe et al., 2017). Methadone’s potential for overdose death (Kaufman et al., 2022) was overridden by its substantial distribution, explaining the underrepresentation (Furst et al., 2022). There has been some prior confusion regarding the safety of methadone relative to its role as the most distributed opioid by MME in the US (Piper et al., 2018). A prior report claimed that methadone accounted for less than five percent of opioid prescriptions dispensed but a third of opioid related deaths (Webster et al., 2011). The data source however, (IMS Health, now known as IQVIA) did not include methadone from the predominant sources of distribution from narcotic treatment programs and other federal programs (Fusca, 2022).

The main strengths of our novel study include the analysis of eleven commonly prescribed opioids, separated into uses for pain and OUD. Limitations of our study involve the ARCOS and FAERS databases. ARCOS does not filter out veterinary uses and there has been drug reimportation data (Bhosle & Balkrishnan 2007). However, the use of opioids among veterinarians was modest (Piper et al., 2020). The FAERS database might have overrepresented select opioids because of duplicates, incomplete results, non-verifiable data, and uncertainty in adverse effect causality (Veronin et al., 2019). The ADEs could also have been underreported since FAERs is a voluntary database submitted by healthcare professionals and others. In this case, most of the opioid ADEs reports were mainly from patients with a one-third from medical professionals which may contribute to heterogenous quality of reports because of differing report behaviors between healthcare professionals and customers (Toki & Ono 2020). Furthermore, since ARCOS does not differentiate formulations and generic vs brand names for the opioids, the proportion of ADEs and opioid counts may need to be further investigated.

Our analysis on the FAERS and ARCOS databases demonstrated general increases in ADEs relative to opioid counts for select opioids with varied relative distributions for their individual ADEs and numbers. This pattern informs us the need for continuous efforts to address the ADEs of specific opioids to inform healthcare policies and change the perspectives of healthcare providers on these drugs and their prescription practices.

## Supporting information

Raw data calculation for figures

## Data Availability

All data produced in the present study are available upon reasonable request to the authors.

---

**Supplementary Figure 1.**
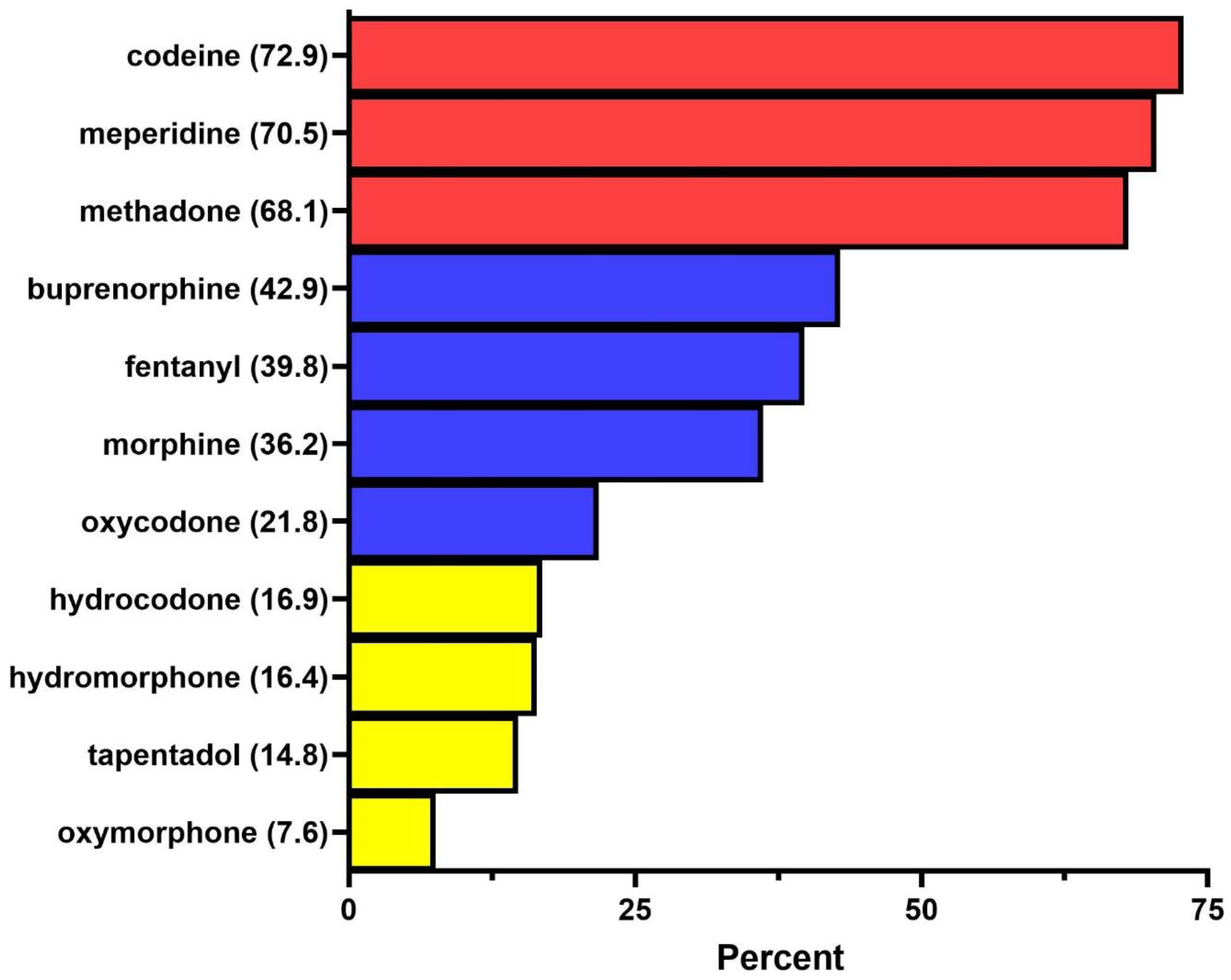
Percentage of reports of eleven opioids reported by healthcare professionals through the US Food and Drug Administration’s Adverse Events Reporting System from 2006-2021.

## References

1. Bhosle, M. J., & Balkrishnan, R. (2007). Drug reimportation practices in the United States. Therapeutics and Clinical Risk Management, 3(1), 41–46.

2. Bishop-Freeman, S. C., Friederich, L. W., Feaster, M. S., & Hudson, J. S. (2021). Buprenorphine-Related Deaths in North Carolina from 2010 to 2018. Journal of Analytical Toxicology, 45(8), 780–791. https://doi.org/10.1093/jat/bkab073

3. Boyle, J. M., McCall, K. L., Nichols, S. D., & Piper, B. J. (2021). Declines and pronounced regional disparities in meperidine use in the United States. Pharmacology Research & Perspectives, 9(4), e00809. https://doi.org/10.1002/prp2.809

4. Burns, R. M., Pacula, R. L., Bauhoff, S., Gordon, A. J., Hendrikson, H., Leslie, D. L., & Stein, B. D. (2016). Policies Related to Opioid Agonist Therapy for Opioid Use Disorders: The Evolution of State Policies from 2004 to 2013. Substance Abuse, 37(1), 63–69. https://doi.org/10.1080/08897077.2015.1080208

5. Cabrera, F. F., Gamarra, E. R., Garcia, T. E., Littlejohn, A. D., Chinga, P. A., Pinentel-Morillo, L. D., Tirado, J. R., Chung, D. Y., Pande, L. J., McCall, K. L., Nichols, S. D., & Piper, B. J. (2019). Opioid distribution trends (2006–2017) in the US Territories. PeerJ, 7, e6272. https://doi.org/10.7717/peerj.6272

6. Centers for Disease Control and Prevention. (2021). Understanding the Epidemic. https://www.cdc.gov/drugoverdose/epidemic/index.html

7. Cicero, T. J., Ellis, M. S., Surratt, H. L., & Kurtz, S. P. (2014). Factors contributing to the rise of buprenorphine misuse: 2008–2013. Drug and Alcohol Dependence, 142, 98–104. https://doi.org/10.1016/j.drugalcdep.2014.06.005

8. Collins, L. K., Pande, L. J., Chung, D. Y., Nichols, S. D., McCall, K. L., & Piper, B. J. (2019). Trends in the medical supply of fentanyl and fentanyl analogues: United States, 2006 to 2017. Preventive Medicine, 123, 95–100. https://doi.org/10.1016/j.ypmed.2019.02.017

9. Comer, S. D., & Cahill, C. M. (2019). Fentanyl: Receptor pharmacology, abuse potential, and implications for treatment. Neuroscience and Biobehavioral Reviews, 106, 49–57. https://doi.org/10.1016/j.neubiorev.2018.12.005

10. Davis, C. S., & Lieberman, A. J. (2021). Laws limiting prescribing and dispensing of opioids in the United States, 1989-2019. Addiction, 116(7), 1817–1827. https://doi.org/10.1111/add.15359

11. Eidbo, S. A., Kropp Lopez, A. K., Hagedorn, J. D., Mathew, V., Kaufman, D. E., Nichols, S. D., McCall, K. L., & Piper, B. J. (2022). Declines and regional variation in opioid distribution by U.S. hospitals. PAIN, 163(6), 1186–1192. https://doi.org/10.1097/j.pain.0000000000002473

12. Fusca E. Accessed 10/9/2022 at: https://pubpeer.com/publications/5B4900548731AB99B907AAAC4AD938

13. Furst, J. A., Mynarski, N. J., McCall, K. L., & Piper, B. J. (2022). Pronounced Regional Disparities in United States Methadone Distribution. Annals of Pharmacotherapy, 56(3), 271–279. https://doi.org/10.1177/10600280211028262

14. Guy, G. P. (2017). Vital Signs: Changes in Opioid Prescribing in the United States, 2006– 2015. MMWR. Morbidity and Mortality Weekly Report, 66(26), 697–704. https://doi.org/10.15585/mmwr.mm6626a4

15. Harrison, L.R.; Arnet, R.E.; Ramos, A.S.; Chinga, P.A.; Anthony, T.R.; Boyle, J.M.; McCall, K.L.; Nichols, S.D.; Piper, B.J. Pronounced Declines in Meperidine in the US: Is the End Imminent?. Preprints 2022, 2022090194 (doi: 10.20944/preprints202209.0194.v1).

16. Johnson, H., Paulozzi, L., Porucznik, C., et al. (2014). Decline in Drug Overdose Deaths After State Policy Changes – Florida, 2010-2012. Morbidity and Mortality Weekly Report, 63(26), 569–574. https://www.cdc.gov/mmwr/preview/mmwrhtml/mm6326a3.htm

17. Kaufman, D.E., Kennalley, A.L., Piper, B.J. (2022). Methadone overdoses increased 48% during the COVID-19 epidemic. medRxiv. https://doi.org/10.1101/2022.04.14.22273870

18. Listos, J., Łupina, M., Talarek, S., Mazur, A., Orzelska-Górka, J., & Kotlińska, J. (2019). The Mechanisms Involved in Morphine Addiction: An Overview. International Journal of Molecular Sciences, 20(17), 4302. https://doi.org/10.3390/ijms20174302

19. Lowe A, Hamilton M MHSc JGBs, Ma J, Dhalla I, Persaud N. (2017). Fatal overdoses involving hydromorphone and morphine among inpatients: a case series. Canadian Medical Association Open Access Journal, 5(1), 184–189. doi:10.9778/cmajo.20160013

20. Lyden, J., & Binswanger, I. A. (2019). The United States opioid epidemic. Seminars in Perinatology, 43(3), 123–131. https://doi.org/10.1053/j.semperi.2019.01.001

21. Mack, K. A., Jones, C. M., & McClure, R. J. (2018). Physician Dispensing of Oxycodone and Other Commonly Used Opioids, 2000–2015, United States. Pain Medicine, 19(5), 990–996. https://doi.org/10.1093/pm/pnx007

22. Mattick, R. P., Breen, C., Kimber, J., & Davoli, M. (2014). Buprenorphine maintenance versus placebo or methadone maintenance for opioid dependence. The Cochrane Database of Systematic Reviews, 2, CD002207. https://doi.org/10.1002/14651858.CD002207.pub4

23. Modarai, F., Mack, K., Hicks, P., Benoit, S., Park, S., Jones, C., Proescholdbell, S., Ising, A., & Paulozzi, L. (2013). Relationship of opioid prescription sales and overdoses, North Carolina. Drug and Alcohol Dependence, 132(1–2), 81–86. https://doi.org/10.1016/j.drugalcdep.2013.01.006

24. Moradi, M., Esmaeili, S., Shoar, S., & Safari, S. (2012). Use of oxycodone in pain management. Anesthesiology and Pain Medicine, 1(4), 262–264. https://doi.org/10.5812/aapm.4529

25. National Institute on Drug Abuse (NIH). (2021 June). Fentanyl DrugFacts. National Institute on Drug Abuse. https://nida.nih.gov/publications/drugfacts/fentanyl

26. O’Donnell, J. K., Gladden, R. M., & Seth, P. (2017). Trends in Deaths Involving Heroin and Synthetic Opioids Excluding Methadone, and Law Enforcement Drug Product Reports, by Census Region—United States, 2006–2015. Morbidity and Mortality Weekly Report, 66(34), 897–903. https://doi.org/10.15585/mmwr.mm6634a2

27. Pashmineh Azar, A. R., Cruz-Mullane, A., Podd, J. C., Lam, W. S., Kaleem, S. H., Lockard, L. B., Mandel, M. R., Chung, D. Y., Simoyan, O. M., Davis, C. S., Nichols, S. D., McCall, K. L., & Piper, B. J. (2020). Rise and regional disparities in buprenorphine utilization in the United States. Pharmacoepidemiology and Drug Safety, 29(6), 708–715. https://doi.org/10.1002/pds.4984

28. Piper, B.J., McCall K.L., Kegan L.R., et al. (2020). Assessment of Controlled Substance Distribution to U.S. Veterinary Teaching Institutions From 2006 to 2019. Frontiers in Veterinary Science, 7:615646. https://doi.org/10.3389/fvets.2020.615646

29. Piper, B.J., Shah, D. T., Simoyan, O. M., McCall, K. L., & Nichols, S. D. (2018). Trends in Medical Use of Opioids in the U.S., 2006–2016. American Journal of Preventive Medicine, 54(5), 652–660. https://doi.org/10.1016/j.amepre.2018.01.034

30. Rege, S. V., Smith, M., Borek, H. A., & Holstege, C. P. (2021). Opioid Exposures Reported to U.S. Poison Centers. Substance Use & Misuse, 56(8), 1169–1181. https://doi.org/10.1080/10826084.2021.1914101

31. Richards, G.C., Aronson, J.K., Mahtani, K.R., Heneghan, C. British Journal of Pain. (2022). Global, regional, and national consumption of controlled opioids: a cross-sectional study of 214 countries and non-metropolitan territories. British Journal of Pain, 16(1),34–40. https://doi.org/10.1177/20494637211013052

32. Romualdi, P., Grilli, M., Canonico, P. L., Collino, M., & Dickenson, A. H. (2019). Pharmacological rationale for tapentadol therapy: A review of new evidence. Journal of Pain Research, 12, 1513–1520. https://doi.org/10.2147/JPR.S190160

33. Seth, P. (2018). Overdose Deaths Involving Opioids, Cocaine, and Psychostimulants— United States, 2015–2016. MMWR. Morbidity and Mortality Weekly Report, 67. https://doi.org/10.15585/mmwr.mm6712a1

34. Singh, G. K., Kim, I. E., Girmay, M., Perry, C., Daus, G. P., Vedamuthu, I. P., De Los Reyes, A. A., Ramey, C. T., Martin, E. K., & Allender, M. (2019). Opioid Epidemic in the United States: Empirical Trends, and A Literature Review of Social Determinants and Epidemiological, Pain Management, and Treatment Patterns. International Journal of Maternal and Child Health and AIDS, 8(2), 89–100. https://doi.org/10.21106/ijma.284

35. Toki T., Ono S. (2020). Assessment of factors associated with completeness of spontaneous adverse event reporting in the United States: A comparison between consumer reports and healthcare professional reports. Journal of Clinical Pharmacy and Therapeutics, 45(3), 462–469. https://doi.org/10.1111/jcpt.13086

36. U.S. Food and Drug Administration (FDA). (2022). FDA Adverse Events Reporting System (FAERS) Public Dashboard. https://fis.fda.gov/sense/app/95239e26-e0be-42d9-a960-9a5f7f1c25ee/sheet/7a47a261-d58b-4203-a8aa-6d3021737452/state/analysis

37. United States Drug Enforcement Administration (DEA). (2018). Drug Scheduling. https://www.dea.gov/drug-information/drug-scheduling.

38. United States Drug Enforcement Administration (DEA). (2022). Division Control Division. ARCOS Retail Drug Summary Reports. https://www.deadiversion.usdoj.gov/arcos/retail_drug_summary/index.html

39. United States Drug Enforcement Administration (DEA). (2019). Diversion Control Division. Drug & Chemical Evaluation Section. Oxymorphone. https://www.deadiversion.usdoj.gov/drug_chem_info/oxymorphone.pdf

40. United States Drug Enforcement Administration (DEA). Intelligence Brief. (2016). Counterfeit Prescriptions Pills Containing Fentanyls: A Global Threat. https://www.dea.gov/sites/default/files/docs/Counterfeit%2520Prescription%2520Pills.pdf.

41. Webster, L. R., Cochella, S., Dasgupta, N., Fakata, K. L., Fine, P. G., Fishman, S. M., Grey, T., Johnson, E. M., Lee, L. K., Passik, S. D., Peppin, J., Porucznik, C. A., Ray, A., Schnoll, S. H., Stieg, R. L., & Wakeland, W. (2011). An Analysis of the Root Causes for Opioid-Related Overdose Deaths in the United States. Pain Medicine, 12(suppl_2), S26–S35. https://doi.org/10.1111/j.1526-4637.2011.01134.x

42. Veronin, M. A., Schumaker, R. P., Dixit, R. R., & Elath, H. (2019). Opioids and frequency counts in the US Food and Drug Administration Adverse Event Reporting System (FAERS) database: A quantitative view of the epidemic. Drug, Healthcare and Patient Safety, 11, 65–70. https://doi.org/10.2147/DHPS.S214771

43. Zhou, Z., & Hultgren, K. E. (2020). Complementing the US Food and Drug Administration Adverse Event Reporting System With Adverse Drug Reaction Reporting From Social Media: Comparative Analysis. JMIR Public Health and Surveillance, 6(3), e19266. https://doi.org/10.2196/19266

